# Remote Neuroinflammation in Newly Diagnosed Glioblastoma Correlates with Unfavorable Clinical Outcome

**DOI:** 10.1101/2024.04.23.24305825

**Authors:** Laura M Bartos, Stefanie Quach, Valerio Zenatti, Sabrina V Kirchleitner, Jens Blobner, Karin Wind-Mark, Zeynep Ilgin Kolabas, Selin Ulukaya, Adrien Holzgreve, Viktoria C Ruf, Lea H Kunze, Sebastian T Kunte, Leonie Hoermann, Marlies Härtel, Ha Eun Park, Mattes Groß, Nicolai Franzmeier, Artem Zatcepin, Adrian Zounek, Lena Kaiser, Markus J Riemenschneider, Robert Perneczky, Boris-Stephan Rauchmann, Sophia Stöcklein, Sibylle Ziegler, Jochen Herms, Ali Ertürk, Joerg C Tonn, Niklas Thon, Louisa von Baumgarten, Matthias Prestel, Sabina Tahirovic, Nathalie L Albert, Matthias Brendel

**Author notes:** **Corresponding author:** Prof. Dr. med. Matthias Brendel Department of Nuclear Medicine, University Hospital of Munich, Marchioninistr.15, 81377 Munich, Germany. **Conflict of interest**: NLA and MB are members of the Neuroimaging Committee of the EANM. JCT received research grants from Novocure and Munich Surgical Imaging and a speaker honorarium from Seagen. NLA received funding from Novocure. MB received speaker honoraria from Roche, GE healthcare and Life Molecular Imaging and is an advisor of Life Molecular Imaging. VCR received speaker honoraria from Novocure. All other authors have declared that no conflict of interest exists. **First author:** Laura Maria Bartos, Department of Nuclear Medicine, University Hospital of Munich, Marchioninistr.15, 81377 Munich, Germany.

## Abstract

Local therapy strategies still provide only limited success in the treatment of glioblastoma, the most frequent primary brain tumor in adults, indicating global involvement of the brain in this fatal disease. To study the impact of neuroinflammation distant of the primary tumor site on the clinical course of patients with glioblastoma, we performed translocator protein (TSPO)-PET in patients with newly diagnosed glioblastoma, glioma WHO 2 and healthy controls and compared signals of the non-lesion (i.e. contralateral) hemisphere. Back-translation in syngeneic glioblastoma mice was used to characterize PET alterations on a cellular level. Ultimately, multiplex gene expression analyses served to profile immune cells in remote brain. Our study revealed elevated TSPO-PET signals in contralateral hemispheres of patients with newly diagnosed glioblastoma compared to healthy controls. Contralateral TSPO was associated with persisting epilepsy and short survival independent of the tumor phenotype. Back-translation pinpointed myeloid cells as the source of TSPO-PET signal increases and revealed a complex immune signature comprised of joint myeloid cell activation and immunosuppression in distant brain regions. In brief, neuroinflammation within the contralateral hemisphere is associated with poor outcome in patients with newly diagnosed glioblastoma. TSPO-PET serves to detect patients with global neuroinflammation who may benefit from immunomodulatory strategies.

## Introduction

Glioblastoma is the most frequent and most aggressive primary brain tumor in adults. The diagnosis is very often associated with high morbidity and poor prognosis. There is a broad body of evidence for the impact of non-neoplastic cells of the tumor microenvironment (TME), such as tumor-associated microglia/macrophages (TAMs), in brain cancer formation, progression (1–4) and treatment response (5, 6). Of note, the abundance of distinct TAM subtypes is reported to independently correlate with overall survival in patients (7, 8) and mice (9) with glioblastoma. Besides, other immune cells such as neutrophils (10–12) and T-cells (13, 14) do also contribute to glioma malignancy and therefore may serve as potential targets for novel therapy approaches. However, the existence of neuroinflammation in brain regions that are not adjacent to the primary tumor site (i.e. remote neuroinflammation) in patients with glioma is only sparsely understood. The possibility to detect tumor (15) and immune cell (16) fragments in liquid biopsies already implies tumor-related changes in the whole body of patients with glioma. An experimental study indicated higher abundance of infiltrating myeloid cells in the contralateral hemisphere of mice with syngeneic glioblastoma (GL26) compared to sham. These infiltrating myeloid cells showed a similar immunosuppressive signature compared to TAMs of the TME (17). In patients, investigations of brain tissue distant to the primary tumor site have so far been limited to autopsy reports (18) that focused on tumor cell infiltration but did not report on neuroinflammation. Hence, our study aimed to investigate neuroinflammation in brain tissue distant of the primary tumor site, as assessed in vivo in patients with glioma, by means of positron emission tomography (PET) of the 18 kDa translocator protein (TSPO) (19). To minimize the impact of tumor infiltration, we focused on the contralateral hemisphere and compared TSPO-PET signals between patients with glioblastoma and healthy controls. We studied dependencies of contralateral TSPO expression from tumor characteristics such as localization, biological tumor volume and TSPO-PET signal magnitude. To investigate potential impact of distant neuroinflammation on patient outcome, we furthermore correlated contralateral PET signal changes with clinical parameters (i.e. epilepsy, overall survival). Back-translation into a glioblastoma mouse model served to pinpoint the cellular source of elevated TSPO-PET signals and to profile the gene expression signature of distant neuroinflammation in glioma.

## Results

### Patient characteristics

A total of 344 patients presenting with glioma that received TSPO-PET imaging were screened (**Fig. 1A**). Out of those, 119 were initial diagnoses, comprising 56 patients with newly diagnosed glioblastoma WHO 4 and eleven patients with newly diagnosed glioma WHO 2 at the time of TSPO-PET. Due to the updated 2021 WHO classification for tumors of the central nervous system (20), two of the selected patients with glioma WHO 2 were excluded to avoid bias due to different therapy regimes (i.e. former WHO 2, isocitrate dehydrogenase (IDH) wild-type) in the following clinical analyses (see below). To allow the investigation of contralateral (non-lesion) hemispheres, patients presenting with bi-hemispherical glioma were excluded. This selection process resulted in 41 patients with glioblastoma, IDH wild-type WHO 4 and nine patients with glioma, IDH mutant (IDHmut) WHO 2 for further TSPO-PET analysis. The characteristics of the cohort are provided in **Table 1**.

**Figure 1:**
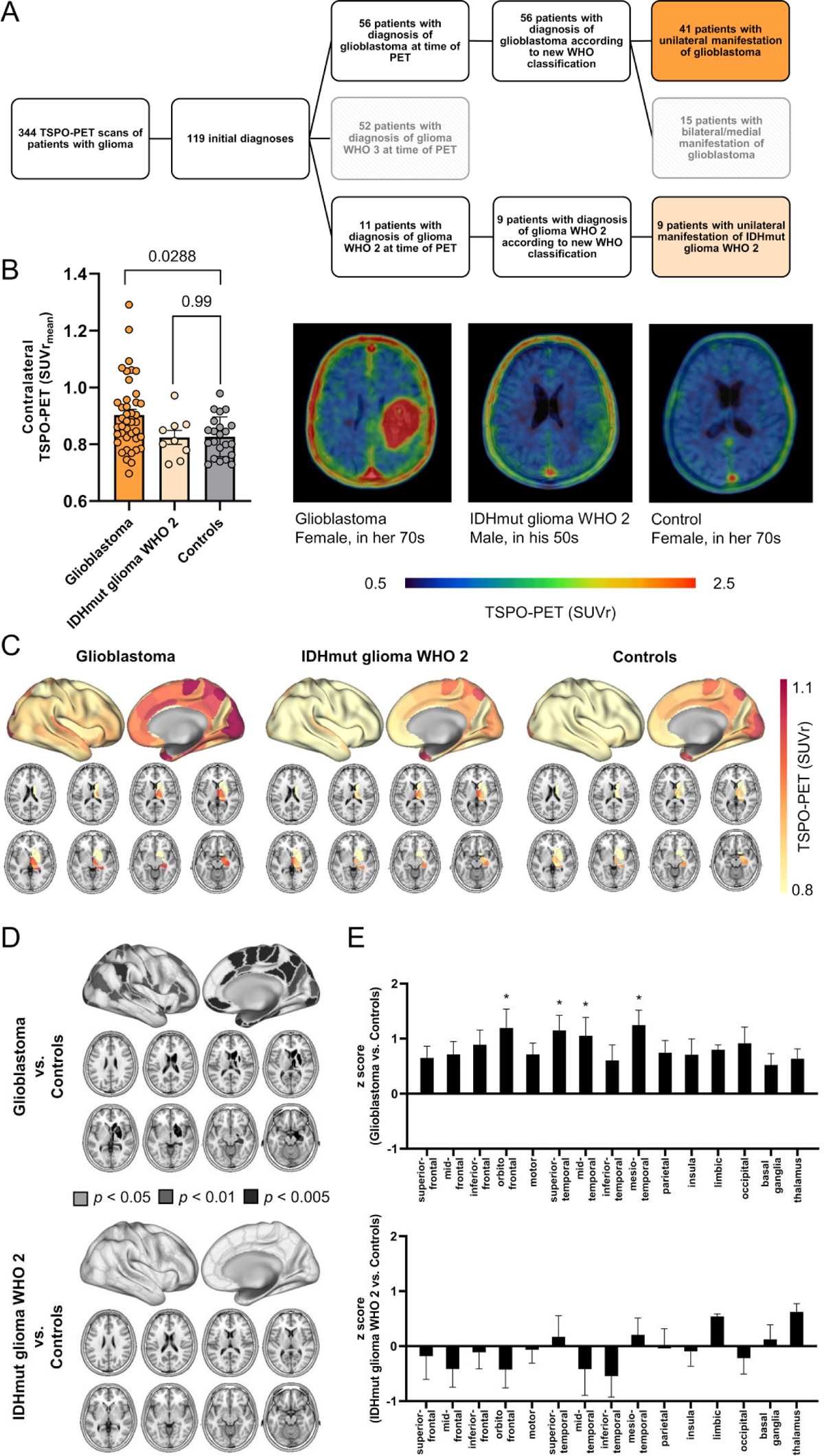
Elevated TSPO-PET signals in the non-lesional hemisphere of patients with glioblastoma. **(A)** Patient selection process. Patients receiving TSPO-PET imaging at inital diagnosis of glioma were allocated by WHO grade. Patients with unilateral manifestation of glioblastoma (WHO °IV; n=41) or Isocitrate dehydrogenase mutant glioma World Health Organization grade 2 (IDHmut glioma WHO 2; n=9) at time of PET were selected for further analysis. **(B)** Patients with newly diagnosed glioblastoma but not patients with IDHmut glioma WHO 2 indicate higher TSPO-PET signal in the contralateral hemisphere compared to healthy controls. Images represent examples of TSPO-PET images of patients with newly diagnosed glioblastoma (left) and IDHmut glioma WHO 2 (middle) in comparison to a healthy control (right). **(C)** Surface projections and axial slices of the group average contralateral TSPO-PET signal of patients with glioblastoma, IDHmut glioma WHO 2 and healthy controls. The non-lesional hemispheres were parcellated into 123 sub-regions using the Brainnetome Atlas. **(D)** Significant TSPO-PET signal elevation in brain regions of the contralateral hemisphere of patients with glioblastoma, but not with IDHmut glioma WHO 2 compared to healthy controls. SFG=superior-frontal gyrus, MFG=mid-frontal gyrus, IFG=inferior-frontal gyrus, OFG=orbitofrontal gyrus, Motor=motor area, STG=superior-temporal gyrus, MTG=mid-temporal gyrus, ITG=inferior-temporal gyrus, MesTemp=mesio-temporal. **(E)** Pronounced contralateral TSPO expression in orbito-frontal, superior-temporal, mid-temporal and mesio-temporal regions of patients with glioblastoma. *p < 0.01.

**Table 1:** Characteristics of the cohort. IDHmut = Isocitrate dehydrogenase mutant. WHO = World Health Organization. SNP = single nucleotide polymorphism. LAB = low, MAB = medium, HAB = high affinity binding status. N.a. = not available. MGMT = O-6-Methylguanine-DNA methyltransferase. TERT = Telomerase reverse transcriptase.

|  | **Glioblastoma** | **IDHmut glioma WHO 2** | **Healthy controls** |
| --- | --- | --- | --- |
| **Sample size** | 41 | 9 | 20 |
| **Tumor type** |  |  |  |
| Astrocytoma | 0 | 7 (78%) | 0 |
| Oligodendroglioma | 0 | 2 (22%) | 0 |
| Glioblastoma | 41 (100%) | 0 | 0 |
| **Age** |  |  |  |
| Median (range) | 71.4<br>(40.0 - 83.9) | 46.4<br>(20.9 - 68.7) | 69.6<br>(24.3 - 80.7) |
| **Sex** |  |  |  |
| Male | 30 (73%) | 5 (56%) | 9 (45%) |
| Female | 11 (27%) | 4 (44%) | 11 (55%) |
| **SNP** |  |  |  |
| LAB | 6 (15%) | 0 | 0 |
| MAB | 16 (39%) | 6 (67%) | 12 (60%) |
| HAB | 16 (39%) | 2 (22%) | 6 (30%) |
| N.a. | 3 (7%) | 1 (11%) | 2 (10%) |
| **MGMT promoter methylation** |  |  |  |
| **status** |  |  |  |
| Methylated | 16 (39%) | 7 (78%) | - |
| Unmethylated | 24 (59%) | 2 (22%) | - |
| N.a. | 1 (2%) | 0 | - |
| **TERT promoter mutation status** |  |  |  |
| Mutant | 36 (88%) | 3 (33%) | - |
| Wildtype | 4 (10%) | 6 (67%) | - |
| N.a. | 1 (2%) | 0 | - |
| **Therapy regimes** |  |  |  |
| Tumor resection | 11 (27%) | 7 (78%) | - |
| Radiotherapy | 37 (90%) | 4 (44%) | - |
| Chemotherapy | 28 (68%) | 2 (22%) | - |
| N.a. | 0 | 1 (11%) | - |
| **Tumor TSPO-PET positivity** |  |  |  |
| Yes | 41 (100%) | 1 (11%) | 0 |
| No | 0 | 8 (89%) | 20 (100%) |

### Patients with glioblastoma but not with IDHmut glioma WHO 2 indicate elevated TSPO-PET signals in the non-lesion hemisphere

To allow atlas-based analyses of tumor-free hemispheres, in a first step, TSPO-PET images of patients with right-sided brain lesions were flipped. The Brainnetome Atlas (21) was used to extract TSPO-PET signals from the non-lesion brain hemisphere (including cortical and subcortical regions) of each patient, excluding surrounding structures such as skull bones or venous sinuses. Contralateral hemispheres of patients with glioblastoma showed a global increase of the TSPO-PET signal when compared to age- and sex-matched healthy controls (+9%, p = 0.022), whereas patients with IDHmut glioma WHO 2 did not indicate relevant changes in contrast to healthy controls (±0%, p = 0.99; **Fig. 1B, C**). In a linear regression model, no significant impact of age (β = 0.173, p = 0.167), sex (β = −0.099, p = 0.428) or the TSPO single nucleotide polymorphism (SNP, β = −0.233, p = 0.067) on the contralateral TSPO-PET signal could be detected. To study these changes in more depth, contralateral hemispheres of patients with glioblastoma, IDHmut glioma WHO 2 and healthy controls were parcellated into 123 sub-regions of the Brainnetome Atlas. This analysis revealed that patients with glioblastoma showed higher TSPO-PET signals (p < 0.05, FDR-corrected) in 60 out of 123 brain regions of the contralateral hemisphere compared to healthy controls, pronounced in the mesio-temporal lobe (+14%; **Fig. 1C, D**). On the contrary, there was no significant TSPO-PET signal elevation in any region of the contralateral hemispheres of patients with IDHmut glioma WHO 2 (all p > 0.5). Going into more detail, the 123 sub-regions were separated into 15 groups of anatomically and functionally connected brain regions (superior-frontal, mid-frontal, inferior-frontal, orbitofrontal, motor area, superior-temporal, mid-temporal, inferior-temporal, mesio-temporal, parietal, insula, limbic, occipital, basal ganglia, thalamus; **Fig. 1E**). Comparing these 15 groups within patients with glioblastoma and IDHmut glioma WHO 2, we found pronounced signal increases in orbito-frontal, superior-temporal, mid-temporal and mesio-temporal regions (p < 0.01, FDR-corrected).

### TSPO-PET signals of the contralateral hemisphere correlate with the tumor phenotype

In a next step, we studied associations between contralateral signal elevations and tumor characteristics. Since patients with IDHmut glioma WHO 2 did not indicate any relevant contralateral TSPO-PET changes compared to controls, we focused on patients with glioblastoma for subsequent analyses. We correlated mean TSPO-PET signal intensities of the contralateral hemisphere with maximum and mean PET signal intensities of the tumor and found a significant correlation for both comparisons (**Fig. 2A**). Interestingly, the contralateral signal elevation was also strongly associated with the tumor volume as assessed via TSPO-PET (R = 0.601, p = 0.00027) and MRI (contrast enhancement (CE): R = 0.367, p = 0.039; T2 signal alteration: R = 0.518, p = 0.0024; **Fig. 2B, C**), but not amino acid PET ([^18^F]FET; R = 0.376, p = 0.077). Correlations were corrected for age, sex and SNP. A regression model using contralateral TSPO-PET signals as outcome variable and mean tumor TSPO-PET signals as well as tumor TSPO-PET volumes as predictors revealed significant associations of both TSPO-PET-dependent variables (tumor SUVr_mean_: β = 0.624, p < 0.0001; tumor TSPO-PET volume: β = 0.251, p = 0.044), together accounting for 55% of the variance in contralateral TSPO-PET signals of patients with glioblastoma (F(_2,35_) = 23.34, p < 0.0001, R^2^ = 0.571, R^2^_Adjusted_ = 0.547). To study regional associations of tumor localization and respective contralateral signal changes, we applied a tumor seed region analysis. The overall correlation of all 123 sub-regions between both hemispheres already indicated regional dependency of contralateral TSPO-PET signal elevations from the particular tumor localization (R = 0.488, p < 0.0001; **Fig. 2D**). Comparing the 15 previously mentioned anatomically and functionally connected brain regions, we observed particularly high contralateral signal elevation when the tumor hotspot was located in orbito-frontal, mesio-temporal or occipital areas (**Fig. 2E, F**). Notably, many tumor seed localizations showed strongest correlation with contralateral regions of the frontal and temporal lobe (**Fig. 2F**), raising the question of effects of this particular regional involvement in higher TSPO expression on patient symptoms.

**Figure 2:**
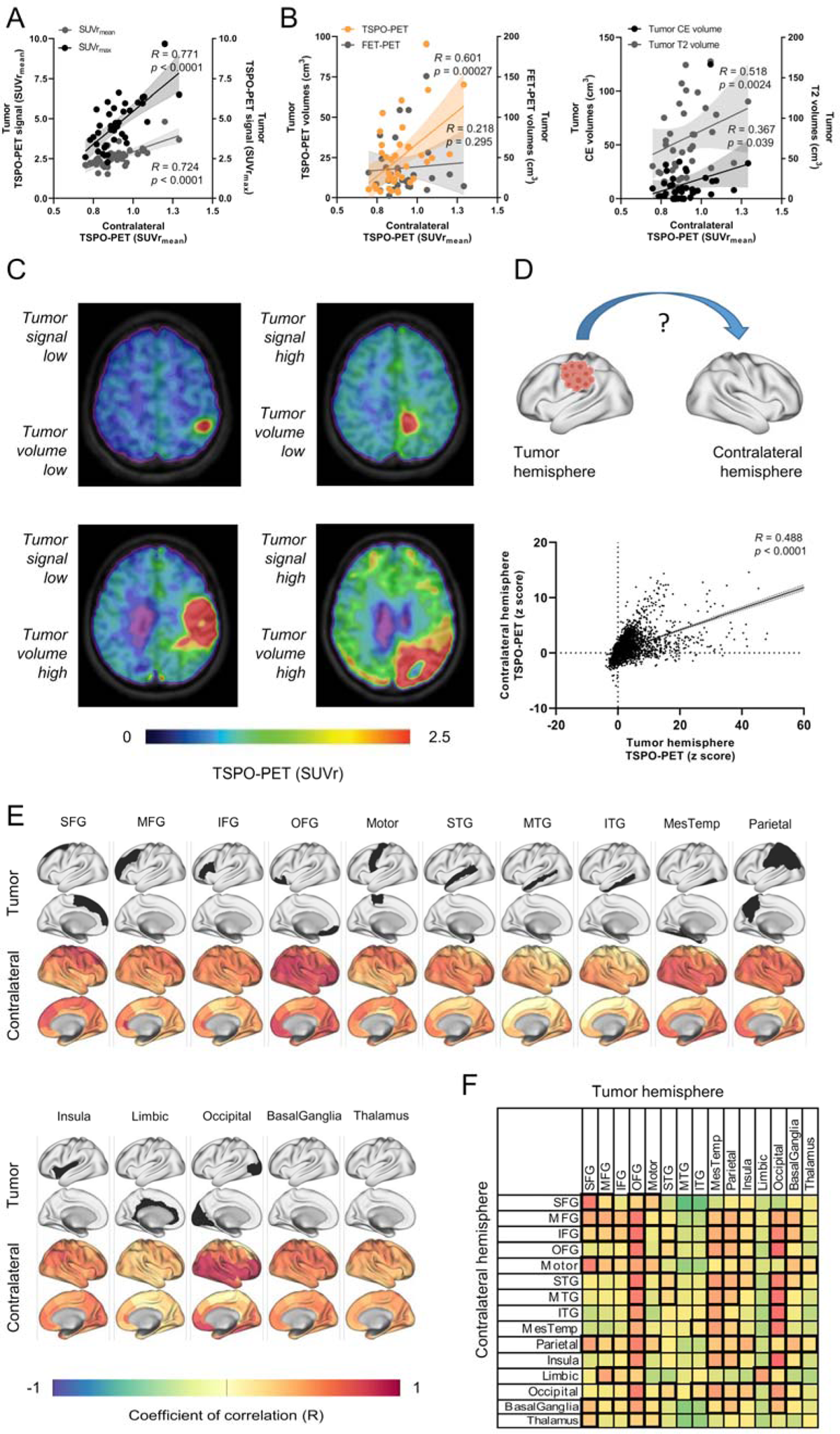
Contralateral TSPO-PET signal elevation is associated with tumor phenotype and localization. **(A, B)** Significant correlation of contralateral TSPO-PET signals in patients with glioblastoma (n=38) with TSPO-PET signal of the tumor (**A**) and the tumor volume in TSPO-PET (**B**, left) and MRI volume (**B**, right). **(C)** Examples of TSPO-PET images derived from patients with distinct tumor extent and TSPO expression in PET indicate distinct contralateral PET signal elevation. **(D)** Overall correlation between corresponding areas of the tumor and the contralateral hemispheres including all 123 brain sub-regions of each individual patient. **(E)** Correlation of regionally defined tumor seed regions with TSPO-PET signals of the 123 sub-regions in the contralateral hemisphere. Surface projection of tumor seed localization (upper rows: seed region, colored in black) and corresponding contralateral coefficients of correlation (lower rows). **(F)** Correlation of both hemispheres within 15 previously defined anatomically and functionally connected brain regions. Highest coefficients of correlation are colored in red, significant correlations are highlighted by thick boundaries.

### Contralateral TSPO-PET signal elevation is associated with persisting epileptic seizures and shorter overall survival in patients with glioblastoma

To study effects of contralateral TSPO-PET signal elevations on patient outcome, we correlated individual contralateral TSPO expression with clinical parameters. Due to pronounced contralateral signal elevation in the temporal lobe, we correlated distant TSPO-PET signals with occurrence of epileptic seizures at initial diagnosis. Although no significant difference could be detected between patients presenting with and without epileptic seizures (p = 0.784, corrected for already initiated glucocorticoid and antiepileptic medication at time of PET), patients with persisting epilepsy after initial successful treatment (i.e. temporary absence of solid tumor) of the primary tumor site showed significantly elevated TSPO-PET signals in the contralateral hemisphere (p = 0.040) compared to patients with discontinued seizures (**Fig. 3A**, additionally corrected for individual tumor therapy regimes). Interestingly, the only two patients presenting with persistent generalized tonic-clonic seizures indicated highest contralateral TSPO-PET signals among the group of patients with glioblastoma suffering from epilepsy (**Fig. 3B**). This finding suggests that epileptic seizures are also associated with alterations in brain regions distant from the primary tumor site. Sub-region analysis revealed elevated contralateral TSPO-PET signals (all p < 0.01) in all temporal areas as well as in frontal and parieto-occipital lobes for patients with glioblastoma and persistent epileptic seizures (**Fig. 3C**). As an exploratory read-out, we investigated regional synchronicity of TSPO-PET within contralateral hemispheres. Here, patients with glioblastoma and persistent epileptic seizures indicated a strong increase of inter-regional TSPO-PET synchronicity compared to patients with discontinued seizures and healthy controls (**Fig. 3D**), speaking for globally elevated TSPO expression in brain as an indicator of epileptic seizures that will not suspend after tumor therapy.

**Figure 3:**
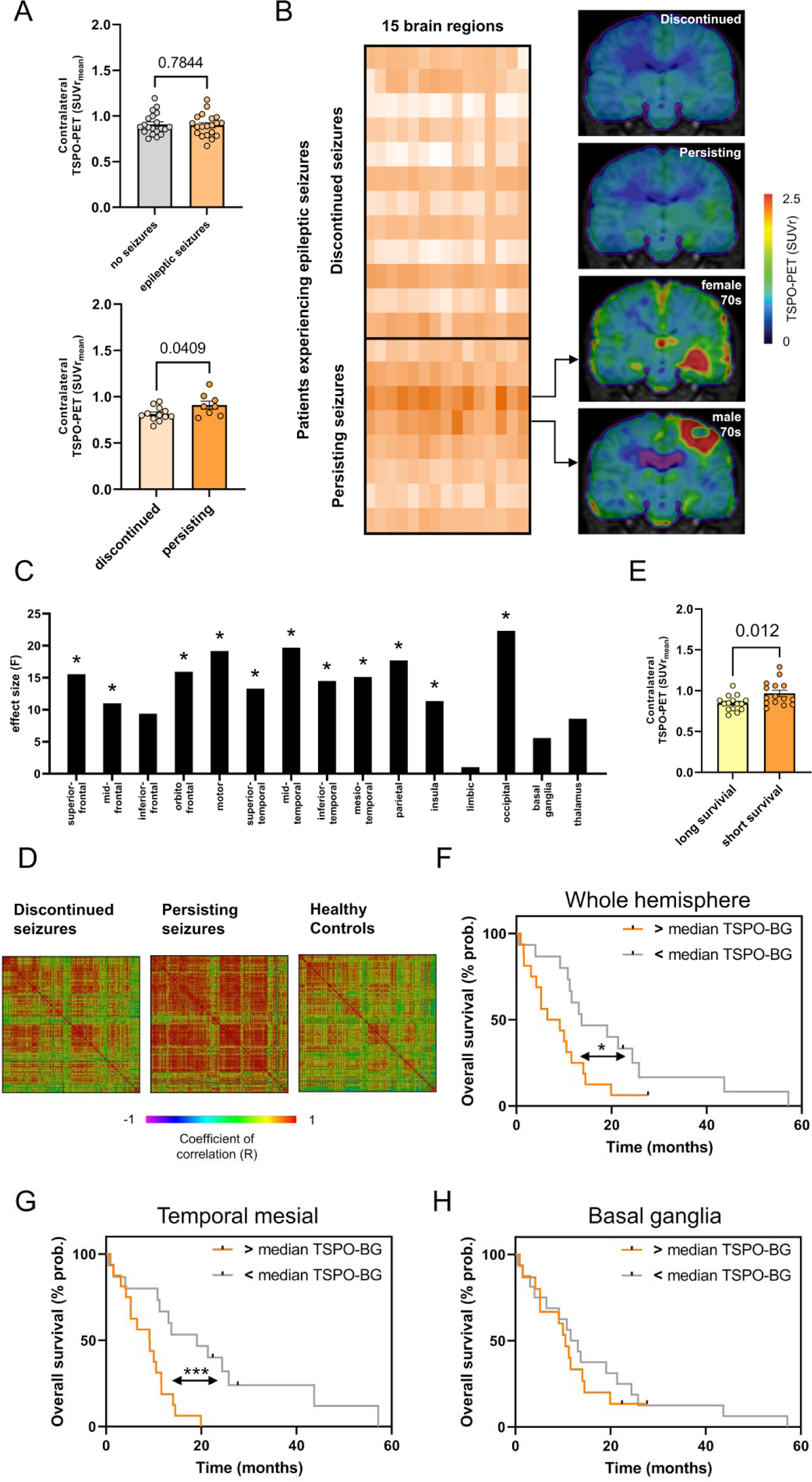
Contralateral TSPO-PET signal elevation is associated with persisting epileptic seizures and worse overall survival. **(A)** Contralateral TSPO-PET shows no quantitative difference between patients presenting with and without epileptic seizures at inital diagnosis (upper panel). Significant increase of contralateral TSPO expression in patients experiencing persisting epileptic seizures after treatment of the primary tumor site compared to patients with discontinued seizures (lower panel). **(B)** Heat map of contralateral TSPO-PET signal in anatomically and functionally predefined brain regions (n=15) of all patients experiencing epileptic seizures at initial diagnosis highlights two patients (age: in their 70s) with strongest contralateral TSPO expression. Coronal slices of TSPO-PET of these two patients are illustrated in comparison to group average images of patients with discontinued and persisting epileptic seizures after tumor therapy. **(C)** Patients with persisting epileptic seizures indicate significant signal elevation in several contralateral brain regions with predominance in motor cortex, mesial temporal lobe and occipital lobe compared to patients with discontinued epileptic seizures. *p < 0.01. **(D)** Matrix of regional TSPO-PET inter-correlation coefficients in comparison of patients with discontinued and persisting seizures as well as controls. Single boxes indicate interregional Pearson’s R. **(E)** Significant increase of the contralateral TSPO-PET signal in patients with short overall survival (≤ 10.7 months, median split). **(F)** High contralateral TSPO-PET signal (> 0.88 SUVr, median split) at initial diagnosis is associated with worse overall survival in patients with glioblastoma. Multivariate Cox regression was adjusted for age, glucocorticoid medication, subsequent radiotherapy and TSPO-PET signal of the tumor. **p < 0.01. **(G)** Distinct predictive value of contralateral TSPO-PET signal in different sub-regions on overall survival. *p < 0.05.

Survival data was available for 31 out of 41 patients with glioblastoma and three patients were still alive at the time of data collection. Patients with glioblastoma and short survival showed higher TSPO-PET signals in the contralateral hemisphere (≤ 11.2 months median survival: SUVr 0.964 ± 0.144) compared to patients with longer survival (> 11.2 months median survival: SUVr 0.847 ± 0.093; F_1,29_ = 7.14, p = 0.012; **Fig. 3E**), also after adjustment for age, sex, the TSPO SNP and initiation of glucocorticoid medication prior to PET as covariates (F_1,25_ = 7.01, p = 0.014). Testing previously evaluated parameters and indices (22) (**Supplemental Table 1**), a significant impact on survival in the current cohort was identified by univariate Cox regression for age (HR: 1.06, p = 0.015), subsequent radiotherapy (HR: 0.27, p = 0.024) and chemotherapy (HR: 0.31, p = 0.003) as well as TSPO-PET signal of the tumor (HR: 1.85, p = 0.024), T1 contrast enhanced volumes (HR: 1.03, p < 0.001) and T2 hyperintensity volumes (HR: 1.02, p < 0.001). Furthermore, high contralateral TSPO-PET signal was associated with shorter survival in univariate Cox regression (HR: 1.72, p < 0.001; Log-rank test after median split: Χ² = 4.22, p = 0.040, **Fig. 3F**). We proceeded with a multivariate Cox regression model including significant indices as well as MGMT, which likely did not reach significance in univariate Cox regression due to the lower sample size compared to a previous study (22), and glucocorticoid medication due to borderline significance. Here, high contralateral TSPO-PET signal was an independent predictor of shorter overall survival within the group of patients with glioblastoma (median OS: 6.5 vs 13.4 months, HR: 2.18, p = 0.005), together with age (HR: 1.12, p < 0.001), MGMT (HR: 0.20, p = 0.010), glucocorticoid medication (HR: 5.16, p = 0.011), and T1 contrast enhanced volume (HR: 1.04, p = 0.025; **Supplemental Table 2**). In terms of validation, high contralateral TSPO-PET signal was still an independent predictor of shorter overall survival (HR: 2.01, p = 0.022) when the whole battery of covariates was included in the multivariate Cox regression (**Supplemental Table 3**). Notably, regions with pronounced TSPO-PET signal elevation in the contralateral hemisphere of patients with glioblastoma compared to controls (**Fig. 1E**) showed stronger impact on overall survival (**Fig. 3G**; i.e. mesial temporal median OS: 9.1 vs 19.1 months, Log-rank test: Χ² = 9.42, p = 0.002) and outperformed regions with lower TSPO elevation (i.e. basal ganglia median OS: 10.5 vs 12.4, Log-rank test: Χ² = 0.38, p = 0.537). In summary, our data indicate dependency of the disease course not only from the local tumor site but also from distant TSPO expression in patients with newly diagnosed glioblastoma.

### scRadiotracing and immunofluorescence identify myeloid cells as the cellular source of altered TSPO-PET signals in the contralateral hemisphere of SB28 glioblastoma mice

To pinpoint underlying sources of contralateral TSPO-PET signal elevations in depth, we back-translated the PET analysis to a SB28 glioblastoma mouse model. Due to lower resolution of murine PET images compared to human, bias due to PET signal spill-over of the tumor into the contralateral hemisphere had to be avoided. Thus, we focused on a sphere of 2 mm diameter placed at a safe distance from the tumor in the frontal lobe of the contralateral hemisphere. SB28 glioblastoma mice indicated higher TSPO-PET signals compared to contralateral hemispheres of sham mice (SUVr: +18%, p = 0.032; VTr: +16%, p = 0.0086; **Fig. 4A, B**). Similar to the human brain, TSPO-PET signals of the tumor correlated with contralateral PET signal enhancement in a voxel-wise analysis (**Fig. 4C**). Next, scRadiotracing (23) was performed in the contralateral hemispheres of SB28 glioblastoma and sham animals with the purpose of allocating the contralateral signal elevation to a certain cell type. Cellular tracer uptake measures identified significantly higher TSPO tracer uptake in myeloid cells of the contralateral hemispheres of SB28 tumor mice compared to sham mice (2.8-fold, p < 0.0001) and healthy control mice (4.2-fold, p < 0.0001; **Fig. 4D**). Again, no significant difference could be detected for sham against healthy mice. Myeloid-cell-depleted fractions of the contralateral hemisphere showed no difference between SB28 tumor, sham and healthy mice (all p > 0.5), underlining specificity of tumor-related contralateral TSPO-PET changes to the myeloid cell population (**Fig. 4D**). To further characterize remote neuroinflammaton, we performed immunofluorescence in the cortex of the contralateral hemisphere of glioblastoma and sham mice and we analyzed the occupancy of IBA1 and TSPO positive myeloid cells. We observed a 2-fold higher myeloid cell (IBA1 positive) coverage comparing glioblastoma and sham animals (p = 0.012; **Fig. 4E, F**). Additionally, we observed a far higher TSPO expression in IBA1 positive cells of the non-lesion hemisphere of glioblastoma mice compared to sham (5.7-fold, p = 0.0048; **Fig. 4E, F**). Considering scRadiotracing and immunofluorescence data together, our results show that higher TSPO-PET signals do not solely derive from higher myeloid cell density in the contralateral hemisphere of glioblastoma animals, but are predominately caused by higher TSPO expression of single myeloid cells (**Fig. 4E, F**). To estimate the influence of TSPO-expressing tumor cells in brain regions remote to the primary tumor site, we performed qPCR and analyzed eGFP expression in SB28 glioblastoma mice at the tumor and contralateral hemisphere (**Fig. 4G**). Relevant eGFP expression could only be detected in the tumor hemisphere. In contrast, we observed higher TSPO expression in the contralateral hemisphere of SB28 glioblastoma mice compared to sham and healthy mice, suggesting that largely increased TSPO expression does not originate from major infiltration of tumor cells at the contralateral site (**Fig. 4G**). In summary, our back-translation approach clearly identified elevated TSPO expression and tracer uptake of myeloid cells as the cellular correlate of high contralateral TSPO-PET signals in glioblastoma.

**Figure 4:**
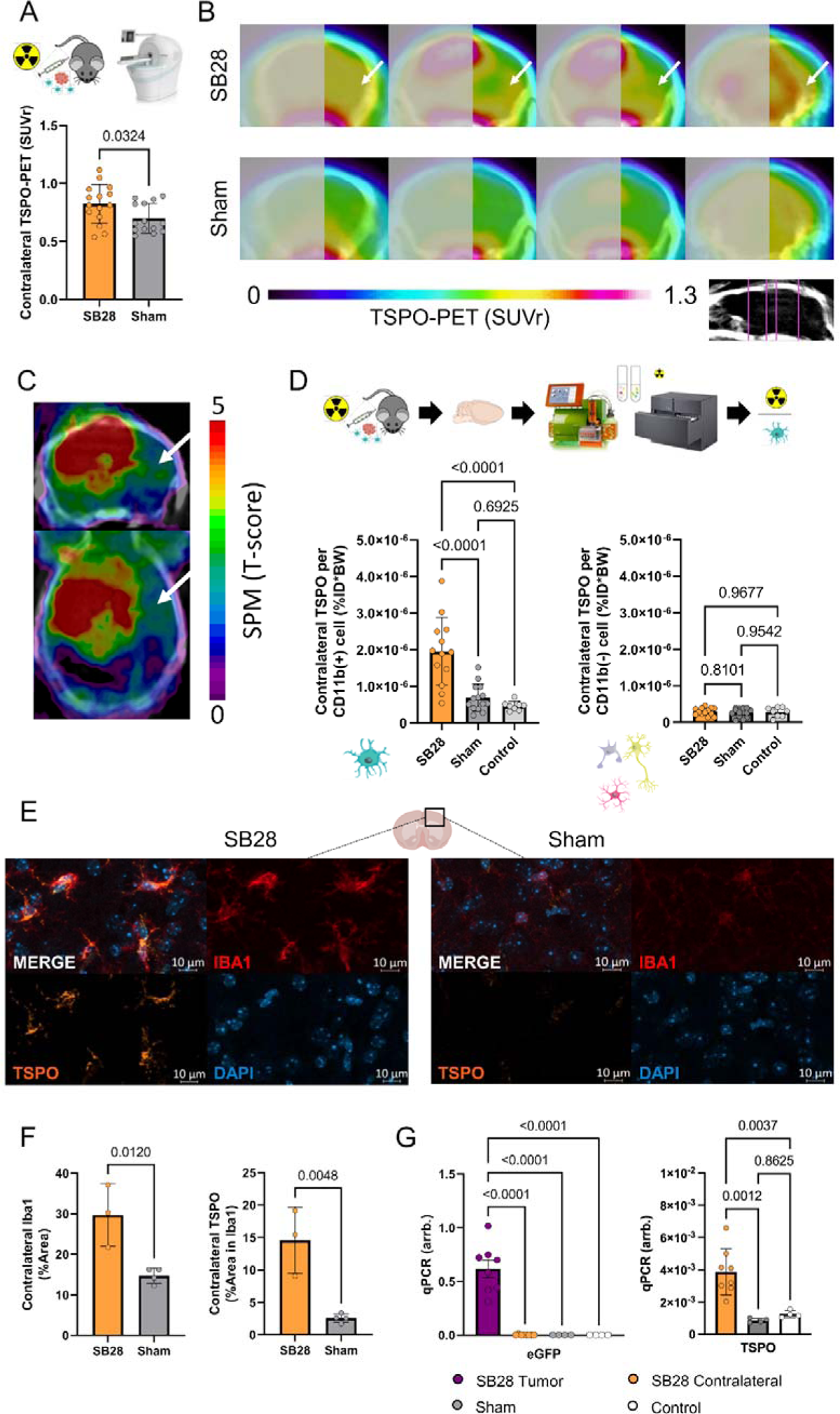
Back-translation into a glioblastoma mouse model pinpoints myeloid cells as the cellular source of contralateral TSPO-PET signal elevations. **(A)** Glioblastoma mice (n=15) show higher TSPO-PET signal in the contralateral hemisphere compared to sham injected animals (n=14). **(B)** Coronal slices of group average TSPO-PET images of SB28 glioblastoma mice (upper row, n=15) and sham injected mice (lower row, n=14). Analyzed planes are indicated upon a CT template. Arrows point to the PET signal elevation in the contralateral hemisphere of SB28 mice. Tumor hemisphere is blurred. **(C)** Whole brain tumor seed correlation analysis shows dependency of contralateral TSPO-PET signals (white arrows) from tumor TSPO-PET signals in SB28 mice (n=15). Tumor TSPO-PET signals were extracted using a spherical volume of interest with 1.5 mm diameter and included as a seed in a voxel-wise statistical parametric mapping (SPM) regression model. **(D)** CD11b-positive myeloid cells of the contralateral hemisphere of glioblastoma mice (n=13) show higher single cell TSPO tracer uptake than myeloid cells of sham injected (n=14) or healthy control mice (n=8, left). Non-myeloid cells show no difference in TSPO tracer uptake between the three conditions (right). **(E, F)** Immunohistochemistry shows higher myeloid cell abundance and increased TSPO in Iba1-positive myeloid cells in contralateral hemispheres of SB28 glioblastoma mice (n=3) compared to sham (n=4). **(G)** qPCR was used to exclude infiltration of SB28 glioblastoma cells into the contralateral hemispheres in SB28 glioblastoma mice (n=8, left). Tumor tissue was used as a positive control. While tumor cells could only be detected in the tumor tissue, TSPO expression was observed in contralateral hemispheres of SB28 glioblastoma mice and exceeded TSPO expression in sham injected (n=4) and healthy control mice (n=4, right).

### Characterization of cells within the contralateral hemisphere of glioblastoma mice shows high expression of genes associated with immune cell activation and immunosuppression

Ultimately, we aimed to characterize the transcriptomic profile of immune cells of the contralateral hemisphere in SB28 glioblastoma mice in depth. We applied the nCounter® Immunology panel and detected significantly higher expression of immune cell markers in the non-lesion hemisphere of glioblastoma animals compared to sham (**Fig. 5A**). In particular, we found strong elevation of key regulators of myeloid cell function, including many disease associated markers, such as CD68, LAMP1/LAMP2, cathepsins, GRN, IBA1 (AIF1), TREM2, TYROBP, FABP5 and TSPO. Increased expression of CD68, PU.1/SPI1 and HEXB was additionally validated using qPCR (**Fig. 5B**). Furthermore, genes associated with immunosuppression of glioblastoma (24) such as CHIL1/CHI3L1 (25) or TGFB1 (26) were increased in distant brain tissue. A pathway analysis revealed 40 out of 50 top rated pathways within the spectrum of cell migration (phagocytes, leukocytes), phagocytosis and immune cell activation (**Fig. 5C**). Ultimately, an IPA network analysis based on altered gene expression signatures in contralateral hemispheres of SB28 glioblastoma mice in contrast to sham reveals TSPO as a mediator for elevated expression of genes associated with poor prognosis (e.g. CCL2 (27)), therapy resistance (e.g. CD74 (28), LCN2 (29)) and immunosuppression (e.g. CHIL1/CHI3L1 (25); **Fig. 5D**), highlighting its potential for in vivo characterization of the immune phenotype in brain of patients with glioblastoma.

**Figure 5:**
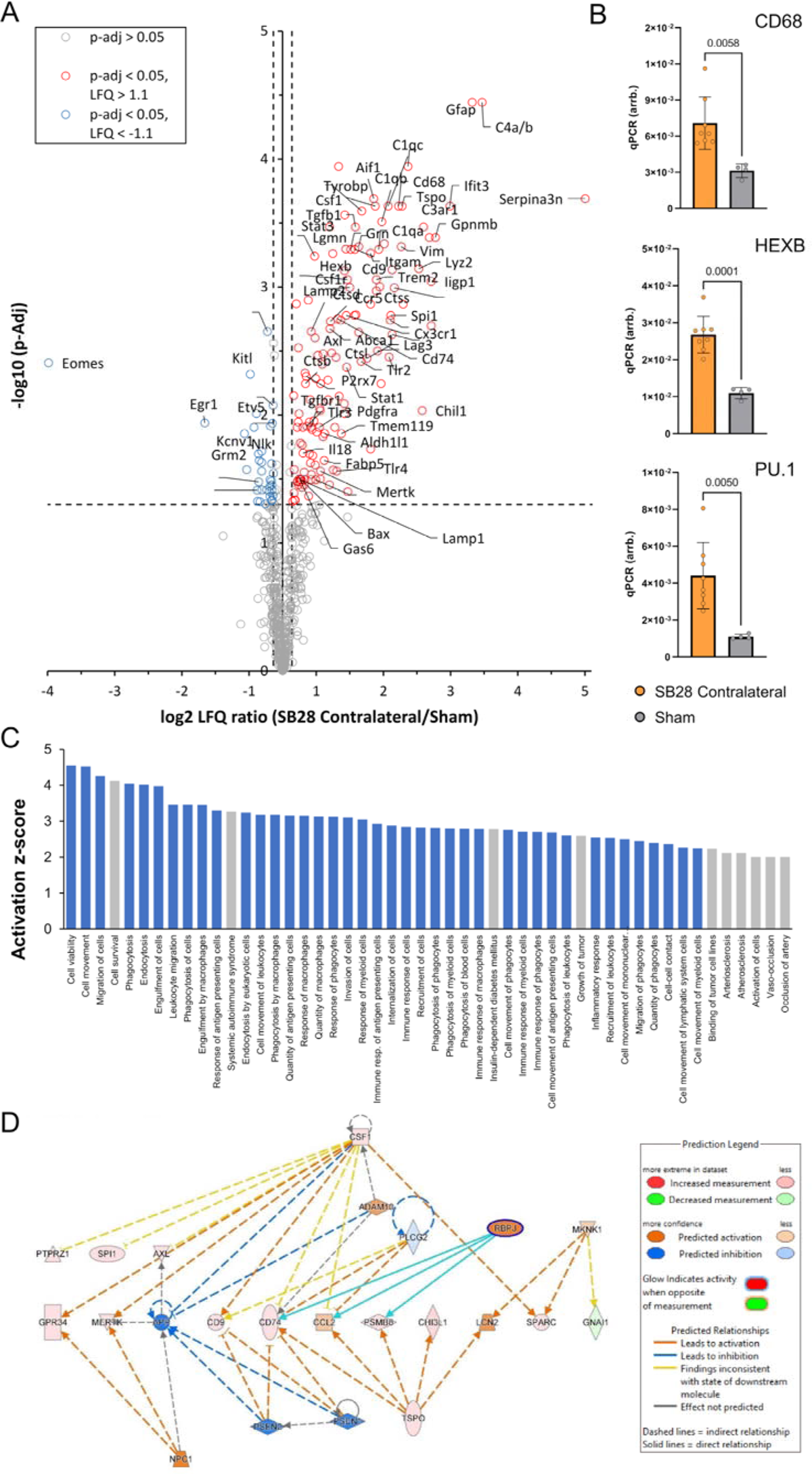
Contralateral hemispheres of SB28 glioblastoma mice indicate signatures of disease associated myeloid cells and immunosuppression. **(A)** Volcano plot highlights genes with significant up- and downregulation in the contralateral hemisphere of SB28 glioblastoma mice (red and blue, respectively) compared to sham. **(B)** qPCR reveals higher expression of genes related to an inflammatory myeloid cell state in contralateral hemispheres of SB28 glioblastoma mice in contrast to sham. **(C)** Illustration of the 50 pathways with highest elevation in immune cells of the contralateral hemisphere of SB28 glioblastoma mice (informed by IPA). Note that 40 out of the 50 pathways with highest activation were related to cell migration (phagocytes, leukocytes), phagocytosis and immune cell activation (blue). **(D)** IPA network analysis based on altered gene expression signatures in contralateral hemispheres of SB28 glioblastoma mice in contrast to sham. Depicted is a network plot showing TSPO-mediated associations with poor prognosis (e.g. CCL2) and immunosuppression (e.g. CHI3L1).

## Discussion

The results of this translational study suggest that neuroinflammation in brain distant of the primary tumor site can be detected in patients with glioma. In particular, TSPO-PET imaging revealed higher contralateral signals in patients with newly diagnosed glioblastoma compared to patients with newly diagnosed IDHmut glioma WHO 2 and healthy controls. Back-translation into a glioblastoma mouse model reflected the increase of contralateral TSPO expression and identified myeloid cells as the source of elevated TSPO-PET signals in distant brain tissue. Remote neuroinflammation in patients with glioblastoma was associated with higher TSPO expression of the tumor and larger tumor extent, also showing regional dependency from the primary tumor site. Most importantly, higher TSPO-PET signals of the contralateral hemisphere were observed in patients with persisting epileptic seizures and associated with shorter overall survival independent of the tumor phenotype.

TSPO was used as a biomarker of glial activation and facilitated non-invasive detection of distant neuroinflammation in living patients with glioblastoma not only compared to controls but also in contrast to patients with IDHmut glioma WHO 2. This main finding of our study was obtained after careful selection of a cohort only consisting of newly diagnosed patients with glioma that underwent TSPO-PET imaging prior to any intervention, since therapy regimes such as radiochemotherapy or surgery cause inflammatory responses in brain (30). Our approach allowed to generate a snapshot of the unmodulated immune response in distant brain tissue containing less sources of bias compared to previous reports on contralateral hemispheres of end-of-life autopsy cases (18). PET imaging served as a valuable tool for biomarker detection in supposedly unaffected tissue of patients with glioma which is not regularly accessible via surgery or biopsy. However, it needs to be acknowledged that PET signal changes only reflect a surrogate of biological alterations at low resolution compared to comprehensive opportunities of tissue workup (i.e. scRNA or multiplex immunohistochemistry). Thus, for critical interpretation of our clinical findings, we performed back-translation into a murine SB28 glioblastoma mouse model and deciphered myeloid cells as the cellular source of increased contralateral TSPO-PET signals. TSPO tracer uptake of the contralateral hemisphere of glioblastoma mice was driven by CD11b-positive myeloid cells with only minor impact of other cell types. Moreover, our preclinical imaging data also confirmed in vivo increases of TSPO expression with dynamic PET imaging quantification that controls for alterations of blood flow.

We note that human TSPO-PET signal changes in tissue distant of the tumor could still be influenced by infiltrating tumor cells, which even exceeded the cellular uptake of the same TSPO tracer compared to myeloid cells (31). To minimize this potential impact of tumor cell invasion on TSPO expression, we focused on the contralateral hemisphere and used only tumors without bilateral manifestation. Thus, sole investigation of the contralateral hemisphere reduced the probability of including direct tumor cell induced immune response in the analysis. However, the occurrence of tumor cell infiltration in the contralateral hemisphere in patients cannot be excluded since autopsy reports and primary human glioblastoma models provided evidence for cross-hemisphere migration of tumor cells along white matter tracts (18, 32–34). Still, contralateral TSPO-PET signal increases, as detected in patients, were robustly observed in the murine SB28 glioblastoma mouse model that only showed appearance of eGFP-positive tumor cells in the ipsilateral hemisphere, which was furthermore validated via qPCR. Moreover, a preclinical study indicated sparing of tumor cell migration into the hippocampal region (35), whereas our human PET analysis showed highest signal elevation in the mesio-temporal lobe. Thus in summary, contralateral changes in TSPO-PET are likely dominated by immune cells within the contralateral hemisphere. Ultimately, our particular assessment of tracer uptake per cell highlighted that elevated TSPO-PET signals in remote brain regions predominantly originated from an altered immune phenotype characterized by increased cellular TSPO expression, with less impact of increased immune cell density.

In this regard, previous preclinical studies reported high contralateral immune cell infiltration in distinct glioblastoma mouse models that additionally show a similar immunosuppressive phenotype compared to TAMs (17, 36). Thus, we asked if similar effects are also present in the contralateral hemisphere of SB28 mice and questioned the significance of TSPO within the glial profile. Strikingly, we found a strong upregulation of genes related to disease associated myeloid cells, such as CD68, LAMP1/ LAMP2, cathepsins, GRN, TREM2, ITGAM, FABP5, TSPO and others that are also often found upregulated in neurodegenerative conditions (37, 38). However, in contrast to downregulation of homeostatic signatures as observed in rodent models of neurodegeneration, we found upregulation of respective genes including HEXB, Mertk, Tmem119, TGFB1 or TGFBR1 in the contralateral hemisphere of SB28 mice. Importantly, some of these genes have already been described as associated with immunosuppression in glioblastoma (24, 26). Key regulators of the myeloid cell lineage such as CX3CR1, CSF1R or PU1/SPI1 were also detected upregulated, reflecting major changes in myeloid gene expression signatures.

TSPO was not only observed as an activator of a variety of pathways associated with poor prognosis, therapy resistance and immunosuppression, but also as one of the genes with highest expression in the contralateral hemisphere of SB28 mice, thus supporting its relevance as a biomarker for monitoring myeloid cell activation (39). From a translational perspective, our findings indicate that TSPO-PET facilitates in vivo assessment of an altered immune cell signature not only in the tumor, but throughout the whole brain of patients with glioblastoma.

In addition to the identification of immune cells as the underlying source of distant TSPO-PET signal increases, we found a spatial dependency of contralateral neuroinflammation from the primary tumor site. In particular, we observed a topological correlation between TSPO-PET signals in tumor seed regions and corresponding contralateral regions in patients with glioblastoma. This indicates that anatomical connections between both hemispheres might be the driver of pronounced contralateral inflammatory response. In this regard, we hypothesize that similar to anterograde tumor cell migration along white matter tracts, these pathways might retrogradely be used by immune cells in brain. Indeed, remote myeloid cell activation along white matter tracts has already been reported in the chronic phase of neuroinflammation after stroke (40). Noteworthy, particular involvement of the frontal and temporal lobe raises the question of associations between pronounced regional contralateral signal elevations and respective symptoms (e.g. frontal lobe syndrome, cognitive dysfunction, epilepsy). Disturbances in functional connectivity, even in the contralateral hemisphere, have already been described for patients with glioblastoma compared to IDHmut glioma (41). Future studies could use the individual functional connectome to interrogate the dependency of contralateral neuroinflammation on brain networks.

Furthermore, we describe a positive correlation of contralateral TSPO-PET signals with tumor volumes in PET and MRI, but also with TSPO expression of the tumor. We speculate that large tumors might cause higher immune response of the whole brain, whereas the positive association between TSPO signal in the tumor and the contralateral hemisphere moreover suggests a gene expression alignment with detrimental immune cell infiltration of the tumor (42) in combination with an immune resistance phenotype (43). This hypothesis was underpinned by the immune panel, showing upregulation of immune resistance genes (24) within contralateral hemispheres of glioblastoma mice. In line, we report significant implications of distant neuroinflammation for clinical patient outcome. First, contralateral TSPO-PET signal increases are predictive for persisting epilepsy after successful treatment (i.e. temporary absence of solid tumor) of the primary tumor site. In line with our findings in glioma, there is also evidence of ipsilateral (44) and contralateral (45, 46) elevation of TSPO in epilepsy, likely representing neuroinflammation. Changes in conventional electroencephalography, also within the contralateral hemisphere, have already been described for patients experiencing acute ischemic stroke (47), additionally reporting an impact of brain alterations distant of the primary lesion site on patient recovery. In this regard, our data also indicate associations of distant neuroinflammation with poor prognosis, even independently from the tumor TSPO expression. Several studies have already described an inflammatory glioblastoma phenotype (48–50), which was shown to be associated with shorter patient survival (51). A recent review on immunotherapy in glioblastoma (52) however suggested that exactly these inflammation-related tumors could be more responsive to immunomodulatory approaches such as immune checkpoint blockade (53) or modulation of novel immune targets e.g. TREM2 (54, 55). Taken together, our findings confirm that glioblastoma should be seen as a whole brain disease and might explain why local therapy approaches have to date not been able to achieve remarkable effects on patient survival. TSPO-PET imaging could serve to detect patients with global neuroinflammation that may profit from immunomodulatory strategies.

## Methods

### Study design

The primary goal of this study was to investigate distant neuroinflammation in the contralateral hemispheres of patients with glioma. For this purpose, we used TSPO as an acknowledged immune PET biomarker in neurological and neuro-oncological diseases and examined contralateral PET signal changes in patients with glioblastoma and patients with IDHmut glioma WHO 2 compared to healthy controls. Patients with IDHmut glioma WHO 2 served as controls with spatially limited disease. Patients presenting with bi-hemispherical glioma were excluded. Contralateral TSPO-PET signal was correlated with tumor PET and MRI indices in patients with glioblastoma. In a next step, the contralateral hemisphere was subsegmented using the Brainnetome Atlas (21) to investigate regional accentuation of contralateral TSPO-PET signal changes including their dependency from tumor localization. Ultimately, associations between contralateral TSPO-PET signal increases and clinical parameters (i.e. epilepsy, overall survival) of patients with glioblastoma were studied.

To disentangle cellular sources of PET signal changes in the contralateral hemisphere of glioblastoma, back-translation into a glioblastoma mouse model was performed. Mice with late-stage syngeneic orthotopic implanted glioblastoma, sham injection, and healthy mice received TSPO-PET imaging for assessment of contralateral signal changes. A combination of radiotracer injection and immunomagnetic cell sorting (scRadiotracing (23)) then served for assessment of TSPO tracer uptake at cellular resolution. In addition, immunofluorescence was performed in mouse brains to investigate myeloid cell occupancy and cellular TSPO expression in contralateral hemispheres of glioblastoma bearing mice compared to sham-injected brains. The mouse model also served to validate the TSPO-PET signal changes observed in humans in absence of tumor cell infiltration to the contralateral hemisphere, since tumor cells can also contribute to tissue TSPO expression (31). Finally, the signature of contralateral immune cells was characterized using the nCounter Immunology panel and qPCR.

### Sex as a biological variable

Our study examined male and female patients and similar findings are reported for both sexes. Human analyses were furthermore adjusted for sex as a covariate. In our preclinical data, only female mice were examined to reduce variability and animals needed in terms of the 3R principle.

### Radiosynthesis

Automated production of [^18^F]GE-180 was performed on a FASTlab^TM^ synthesizer (GE HealthCare) with single-use disposable cassettes. The pre-filled precursor vial was assembled on the cassette and the cassette was mounted on the synthesizer according to the set-up instructions. The FASTlab^TM^ control software prompts were followed to run the cassette test and to start the synthesis. No-carrier-added ^18^F-fluoride was produced via ^18^O(p, n)^18^F reaction by proton irradiation of ^18^O-enriched water and delivered to the ^18^F incoming reservoir. The fully automated manufacturing process consists of the following steps: trapping of ^18^F-fluoride on a QMA cartridge, elution using Kryptofix^®^222, potassium hydrogen carbonate, water and acetonitrile, azeotropic drying of ^18^F-fluoride at 120°C for 9 minutes, labelling of the precursor in MeCN at 100°C for 6 minutes, dilution of the crude product with water, tC18 cartridge based purification by use of 20 mL 40% (v/v) Ethanol and 11.5 mL 35% (v/v) Ethanol, elution of the product with 3.5 mL 55% (v/v) Ethanol and final formulation with phosphate buffer. RCY 39 ± 7% (n = 16) non d. c., synthesis time 43 minutes, RCP ≥ 98%.

### Patients and healthy controls

All patients with suspected glioma who received TSPO-PET at the Department of Nuclear Medicine, University Hospital, LMU Munich between August 2016 and November 2021 were screened for the study. Eligible for analysis were adult patients with [^18^F]GE-180 PET at initial diagnosis of glioma prior to start of therapy (**Fig. 1A**). Patients were stratified into two groups after tissue sample analysis (stereotactic biopsies or tumor resection), extracted at the Department of Neurosurgery, University Hospital, LMU Munich and evaluated at the Center for Neuropathology, LMU Munich. In the first group, we allocated patients with diagnosis of glioblastoma according to histopathological determination. This group contained only patients who were diagnosed with glioblastoma at the time of PET imaging and remained classified as glioblastoma in the current version of the World Health Organization (WHO) classification for tumors of the central nervous system (20). In the second group, we allocated patients with diagnosis of glioma WHO grade 2 at the time of PET imaging that remained classified as glioma WHO grade 2 in the current version of the 2021 WHO classification for tumors of the central nervous system (20). The rationale was to include a comparison group with disease but limited tumor cell infiltration and we did therefore not include patients with glioma WHO grade 3. All other patients were excluded from the analysis to avoid bias due to different therapy regimes (i.e. former WHO grade 2, IDH wild-type). All tumor samples were analyzed regarding their molecular genetics (e.g. wild-type status of the IDH1/2 gene, MGMT promoter methylation, TERT promoter mutation). Genotyping for a single nucleotide polymorphism (SNP) of the TSPO gene was performed at the Department of Psychiatry of the University Hospital Regensburg as described previously using blood samples (56). Accordingly, the patients were categorized as low, medium, or high affinity binders (LAB, MAB, HAB) (57).

### Human PET/CT and MRI acquisition and analysis

All human TSPO-PET scans were performed on a Biograph 64 PET/CT scanner (Siemens). Tracer production, image acquisition and image reconstruction was performed as described previously (58). Approximately 180 MBq [^18^F]GE-180 were injected as an intravenous bolus and summation images 60-80 minutes p.i. were used for image analysis. Magnetic resonance imaging (MRI) sequences included contrast enhanced and native T1- and T2-weighted images. As an additional estimate for tumor volume, O-(2-[18F]-fluoroethyl)-L-tyrosine ([^18^F]FET) PET was performed in a subset of n=28 patients with glioblastoma. Acquisition, reconstruction and analysis of [^18^F]FET-PET were performed as described previously (59).

All TSPO-PET image analyses were performed by PMOD (version 3.5/version 4.3, PMOD Technologies). TSPO-PET images were normalized by scaling to the injected dose (ID) and accounting for body weight (BW) of each patient (standardized uptake values, SUV). For final analysis, images were furthermore intensity-normalized to a spherical volume of interest (VOI) placed in the center of the pons to obtain maps of relative regional TSPO tracer uptake as standardized uptake value ratios (SUVrs). The pons was used as a reference tissue after validation of similar SUV values in that region between all patient cohorts as described before (60).

TSPO-PET SUVr (mean and maximum) of the tumor was acquired from the semi-automatically delineated 3D iso-contouring VOI that also served for TSPO-PET tumor volume assessment as described below. The main analysis strategy consisted of obtaining TSPO-PET SUVr values in n = 246 predefined atlas VOIs, which were used to parcellate the brain into anatomical regions and functional networks by use of the Brainnetome Atlas (21). SUVr values were obtained for all VOIs located in contralateral and ipsilateral hemispheres (n = 123 each).

### Assessment of tumor volumes

MRI and PET segmentation was performed as described previously (59). In brief, images from all three MRI sequences (Contrast enhanced and native T1-weighted MRI, and T2-weighted MRI) were first normalized to white matter intensity as assessed on the contralateral side of the lesion (TBR_CE_, TBR_native_, TBR_T2,_ TBR = tumor-to-background-ratio). The background-normalized CE T1-weighted MRI image was divided voxel-wise by the corresponding background-normalized native T1-weighted MRI image, thus creating relative CE (rCE) images. Since no pre-contrast calibration sequence has been performed, CE MRI intensity normalized to native MRI intensity (i.e., rCE) was assumed to be proportional to the relaxation rate R1. Hence, a linear relationship between contrast agent concentration and rCE was assumed, which is valid for sufficiently small concentrations (61, 62). The ratio rCE between pre- and post-contrast measurements was used as surrogate marker for blood-brain barrier leakage, as applied previously (63). The tumor volumes for rCE, T2 hyperintensities, and TBR_FET_ were defined by including all voxels with abnormal signal in the applied modality, using threshold-based segmentation.

This was achieved by application of the following iso-contour thresholds. For [^18^F]FET the biological tumor volume was defined using the biopsy-proven threshold of 1.6 in TBR_FET_ images (64, 65) which was also used for rCE. Segmentation of hyperintense tumor volumes in TBR_T2_ images was performed using a threshold of 1.3. TSPO-PET thresholds were derived using a line profile assisted visual definition of the tumor border by an expert reader since potential signal changes in the contralateral hemisphere did not provide a rationale to use TBR. TSPO-PET tumor volumes were semi-automatically delineated using a 3D iso-contouring VOI tool with region growing function (version 4.3, PMOD Technologies). In tumors located in vicinity to sinus or calvaria the adjacent structures were cut previous to tumor delineation. Three patients with glioblastoma were not eligible for the 3D analysis of TSPO-PET at the tumor site due to merged signals between tumor and adjacent structures. Semi-automatic segmentation of volumes using the above-mentioned thresholds was based on an initial manual definition of a confining volume containing all tissue areas suspicious in at least one of the modalities, taking into account a wide safety margin that spreads into unaffected tissue, while simultaneously excluding only vessels and healthy ventricles. Within this confining volume, a region growing algorithm was applied for thresholding with an iso-contour starting from user defined seed points. The manual part of the described procedure was performed within the Medical Imaging Interaction Toolkit graphical user interface (MITK; ITK version 4.13.2, VTK version 8.1.0 Qt version 5.12.8 (66, 67)) and the automatic region growing was performed with SimpleITK (version 1.2.4, (68)) using Python 3.8 or PMOD (version 4.3, PMOD Technologies).

### Animals

All experiments were conducted using eight-week-old C57BL/6 mice that were purchased from Charles River (Sulzfeld, Germany) and acclimated for at least 1 week. For TSPO-PET images and scRadiotracing experiments, at day 0, n = 48 mice were inoculated with 100,000 SB28-eGFP cells suspended in 2 µl of DMEM (Merck) (glioblastoma mice, n = 26) or 2 µl of saline (sham mice, n = 22). Additional n = 12 mice received no intervention before scRadiotracing, serving as control animals. For inoculation, mice were anesthetized with intraperitoneal (i.p.) injections of 100 mg/kg ketamine 10% and 10 mg/kg xylazine 2% in 0.9% NaCl. Anesthetized mice were immobilized and mounted onto a stereotactic head holder (David Kopf Instruments) in the flat-skull position. After surface disinfection, the skin of the skull was dissected with a scalpel blade. The scull was carefully drilled with a micromotor high-speed drill (Stoelting) 2 mm posterior and 1 mm left of the bregma. By stereotactic injection, 1 x 10^5^ cells were applied with a 10 µl Hamilton syringe (Hamilton) at a depth of 2 mm below the drill hole. Cells were slowly injected within 1 minute and after a settling period of another 2 minutes the needle was removed in 1 mm steps per minute. After that, the wound was closed by suturing. Mice were checked daily for tumor-related symptoms and sacrificed when tumor burden (i.e. appearance, coordinative deficits, motor symptoms) reached stop criteria (not reached in any animal). On the last day of the experiment, the mice were injected with 13.8 ± 3.1 MBq [^18^F]GE-180 into the tail vein. After PET imaging, cervical dislocation and brain extraction was performed at 75 minutes post injection. N = 15 glioblastoma and n = 14 sham mice received TSPO-PET imaging directly before brain extraction. Out of those, n = 13 glioblastoma and all sham animals were used for scRadiotracing. N = 8 mice without prior intervention served as untreated control animals for scRadiotracing experiments. N = 3 glioblastoma and n = 4 sham-injected mice received perfusion with 4% PFA prior to immunofluorescence. Contralateral hemispheres of n = 8 glioblastoma, n = 4 sham-injected mice and the right hemisphere of n = 4 untreated control mice were snap-frozen in liquid nitrogen prior to immune panel and qPCR analyses. N = 8 snap-frozen tumors served as positive control for eGFP quantification via qPCR.

### Cell culture

SB28-eGFP cells (69) were cultured in DMEM containing MEM non-essential amino acids (1x), 1% Penicillin-Streptomycin solution (Thermo Fisher Scientific) and 10% fetal bovine serum (FBS, Biochrome). Cell cultures were maintained in the incubator at 37°C in humidified and 5% CO_2_-conditioned atmosphere. Cells were passaged when the cell density in the flask reached 80% confluence.

### Small-animal PET/CT acquisition and analysis

All small animal positron emission tomography (PET) procedures followed an established standardized protocol for radiochemistry, acquisition, and post-processing (70, 71). For tumor, sham and healthy control mice, [^18^F]GE-180 TSPO small-animal PET recordings with an emission window of 0-60 minutes after injection were obtained to measure cerebral TSPO expression prior to cell sorting using a Mediso PET/CT system (Mediso). All small-animal PET experiments were performed with isoflurane anesthesia (1.5% at time of tracer injection and during imaging; delivery 3.5 L/min).

All analyses were performed by PMOD (version 3.5, PMOD Technologies) using computed tomography (CT, tumor and sham mice) for spatial normalization (70) and conducted as previously described (72). Radioactivity normalization of PET images was performed analogously to human data (pons scaling; static 40-60 minute frame). Additionally, to acknowledge potential PET signal dependency from perfusion alterations, we calculated volume of distribution ratio (VTr) images with an image derived input function (73) using the methodology described by Logan et al. implemented in PMOD (74). The blood input curve was obtained from a standardized bilateral VOI placed in both carotid arteries (75). A maximum error of 10% and a VT threshold of 0 were selected for modelling of the full dynamic imaging data with a one-tissue compartment model. Time of the linear fit was set flexible (t*) and uptake rate constant K_1_ and dissociation rate constant K_2_ were obtained for the lesion. The obtained VT images were scaled by the pons to generate VTr images.

TSPO-PET SUVr and VTr were obtained from a spherical volume of interest (2 mm diameter) in the lateral frontal pole of the contralateral side of the tumor to avoid spill over from the tumor and adjacent structures of the brain. PET data of the tumor hemisphere have already been published elsewhere (31).

### Immunomagnetic cell separation

MACS was performed as described previously (23, 31, 76). Detailed descriptions of brain dissociation and isolation of myeloid cells were as follows.

### Mouse brain dissociation

Adult mouse brains were removed after cervical dislocation at 75 minutes p.i. and stored in cold D-PBS. The brains were cut into small pieces and dissociated using gentleMACS Octo Dissociator with Heaters (Miltenyi Biotec, 130-096-427) in combination with Adult Brain Dissociation Kit, mouse and rat (Miltenyi Biotec, 130-107-677) for adult mouse brain dissociation of contralateral hemispheres of glioblastoma, sham and healthy control mice. The dissociated cell suspension was applied to pre-wet 70-µm Cell Strainer (Miltenyi Biotec, 130-110-916). The cell pellet was resuspended using cold D-PBS and cold debris removal solution. Cold D-PBS was gently overlaid on the cell suspension and centrifuged at 4°C and 3000 g for 10 minutes with acceleration at 9 and deceleration at 5. The two top phases were removed entirely. The cell pellets were collected. Cell pellets were additionally resuspended with 1 mL of cold red blood cell removal solution followed by 10 minutes incubation. Cell pellets were collected for further applications.

### Isolation of myeloid cells

Myeloid cells were isolated from animals using CD11b MicroBeads, human and mouse (Miltenyi Biotec, 130-049-601) and a MACS separation system (Miltenyi Biotec). The prepared cell pellets were resuspended in 90 µl of D-PBS–0.5% BSA buffer per 10^7^ total cells. Ten microliters of CD11b MicroBeads per 10^7^ total cells were added and incubated for 15 minutes in the dark at 4 °C. Cells were washed by adding 1 mL of buffer per 10^7^ cells and centrifuged at 300 g for 10 minutes. The cell pellets were resuspended in 500 µl of D-PBS– 0.5% BSA. The pre-wet LS columns (Miltenyi Biotec, 130-042-401) were placed onto a QuadroMACS Separator (Miltenyi Biotec, 130-090-976). The cell suspensions were applied onto the column. The columns were washed with 3 × 3 mL of D-PBS–0.5% BSA buffer. The flow-through containing the unlabeled cells was collected as the myeloid-cell-depleted fraction. The columns were removed from the magnetic field, and myeloid cells were flashed out using 5 mL of D-PBS–0.5% BSA buffer.

### Gamma emission measurements

Radioactivity levels of contralateral hemispheres (n = 13 glioblastoma and n = 13 sham-injected mice) and myeloid cell enriched and depleted cell pellets were measured in a gamma counter (Hidex AMG Automatic Gamma Counter), normalized to the injected dose and the body weight, with decay correction to time of tracer injection for final activity calculations.

### Flow cytometry

Flow cytometry staining was performed at 4 °C. The cell suspension was centrifuged at 400 g for 5 minutes and the supernatant was aspirated completely. The cell pellet was then resuspended in 100 µl of cold D-PBS containing fluorochrome-conjugated antibodies recognizing mouse CD11b (Miltenyi Biotec, 130-113-810) in a 1:100 dilution and incubated for 10 minutes at 4 °C in the dark. Samples were washed with 2 mL of D-PBS and centrifuged for 5 minutes at 400 g. Finally, cell pellets were resuspended in 500 μl of D-PBS. Flow cytometry was performed after gamma emission measurement using a MACSQuant® Analyzer. Acquired data included absolute cell numbers and purity of CD11b(+) cells in each sample.

### Calculation of single cell TSPO tracer signal

Measured radioactivity (Becquerel, Bq) of cell pellets was divided by the specific cell number in the pellet resulting in calculated radioactivity per cell. Radioactivity per cell was normalized by injected radioactivity and body weight (i.e. %ID*BW).

### Transcardial perfusion

Mice that were intended for immunofluorescence were i. p. injected with 100 mg/kg ketamine 10% and 10 mg/kg xylazine 2% in 0.9% NaCl. After expiration of the pedal reflex, intracardial perfusion was performed with 0.1 M PBS (10 U/mL, Ratiopharm) for 6 minutes, followed by the administration of 4% paraformaldehyde (PFA) in 0.1 M PBS (pH 7.4) (Morphisto, 11762.01000) for another 6 minutes. Afterwards, the brain was removed and post-fixed by 4% PFA for 4-6 hours at 4 °C, then washed with 0.1 M PBS and stored in 0.1 M PBS.

### Immunofluorescence

30 µm coronal brain sections were obtained from n = 3 SB28 and n = 4 sham mice. The sections were first permeabilized for 30 minutes with 0.5% Triton diluted in PBS (PBS-T) and were incubated for 1 hour in blocking solution (5% normal goat serum/0.5%Triton X-100 in PBS). Then, the sections were incubated with primary antibody diluted in blocking solution at 4 °C for 16 hours. The following primary antibodies were used: rabbit anti-TSPO (ab109497, abcam, 1:500), guinea pig anti-Iba1 (234308, Synaptic Systems, 1:500). The next day, the sections were washed 3 times in PBS-T and incubated with secondary antibodies (Alexa Fluor, Invitrogen, 1:500) and Hoechst (1:2000, Invitrogen) diluted in blocking solution for 2 hours at room temperature. After three washes with PBS-T and one with PBS, the sections were mounted on glass slides using Fluoromount (Sigma-Aldrich).

### Image analysis

Detailed and tile scan images were acquired with using a LSM900 confocal microscopy (Zeiss). Signal coverage for Iba1 was quantified using ImageJ. Furthermore, TSPO Area% was obtained in thresholded Iba1-positive areas.

### qPCR

SB28 glioblastoma brain was acutely isolated from the animal and sectioned in the two hemispheres and the tumor area. Each portion was snap frozen in liquid nitrogen and pulverized using a dry pulveriser (Covaris). RNA was extracted from 10 mg of brain powder using a RNeasy mini kit (Qiagen) and concentration and quality control was assessed using an Infinite 200 PRO plate reader (Tecan). RNA was retrotranscribed using M-MLV Reverse Transcriptase (Promega) and random nonamer primers (Sigma-Aldrich) for 1 hour at 37°C. Gene expression was quantified using a QuantStudio™ 5 Real-Time PCR System (Applied Biosystems) and the following TaqMan Real-Time-PCR-Assay probes: Aif1 (Mm00479862_g1), CD68 (Mm03047343_m1), Clec7a (Mm01183349_m1), eGFP (Mr04329676_mr), Hexb (Mm01282432_m1), Spi1 (Mm00488140_m1), Tspo (Mm00437828_m1). Gene expression was normalized to actin expression (Mm02619580_g1).

### Nanostring

Snap frozen non-lesion hemispheres were pulverized using a dry pulveriser (Covaris). RNA was extracted from 10 mg of brain powder using a RNeasy mini kit (Qiagen) and concentration and quality control were assessed with Infinite 200 PRO plate reader (Tecan) and Bioanalyzer 2100 (Agilent). For all samples RIN values were above 8. Gene expression analysis was performed with nCounter Analysis System (Nanostring). 5 µl of 20 ng/µl dilutions of RNAs extracted were analyzed according to the manufacturer’s protocol. Data were further analyzed with the use of QIAGEN IPA (QIAGEN Inc., https://digitalinsights.qiagen.com/IPA)(77). Standard settings were used for the analysis.

### Statistics

Unless otherwise specified, graphs show mean values ± standard deviation. A p-value < 0.05 was considered significant.

#### Assessment of contralateral TSPO-PET signal in patients

One-way ANOVA was used for comparison of contralateral TSPO-PET signals between patients with glioblastoma and IDHmut glioma WHO 2 and healthy controls. Simple linear regression was used to determine the impact of age, sex or the TSPO SNP on the contralateral TSPO-PET signal. Unpaired Student’s t-test served to compare TSPO-PET SUVrs and z scores of different brain regions in the contralateral hemisphere between study groups. False Discovery Rate (FDR) correction was applied for multiple comparisons.

#### Association of contralateral TSPO-PET signals with tumor phenotype

Three patients with glioblastoma were not eligible for the 3D analysis of TSPO-PET at the tumor site due to merged signals between tumor and adjacent structures. Partial correlation with correction for age, sex and the TSPO SNP was applied for associations of contralateral TSPO-PET signal elevations with the individual tumor phenotype. Multiple regression served for determination of the impact of TSPO-PET-dependent variables on contralateral TSPO-PET signals. Simple linear regression was used for the inter-correlation between the tumor and non-lesion hemisphere. Bonferroni correction was applied for multiple comparisons.

#### Impact of contralateral TSPO-PET signal elevation on patient outcome

One patient with glioblastoma was excluded for correlation of contralateral TSPO-PET signals with occurrence of epileptic seizures due to secondary epilepsy after ischemic stroke prior to tumor diagnosis. Stroke occurred in the hemisphere with future tumor occurrence. Unpaired Student’s t-test was used for comparison of contralateral TSPO-PET signals between patients with and without epileptic seizures (corrected for glucocorticoid and antiepileptic medication at time of PET), and between patients with discontinued and persisting epileptic seizures (additionally corrected for therapy regimes i.e. tumor resection, chemotherapy, radiotherapy). Heat map depicts individual regional z scores of patients with glioblastoma experiencing epileptic seizures. Effect sizes of different brain regions in the contralateral hemisphere were compared via one-way ANOVA. FDR-correction was applied for multiple comparisons. Connectivity maps of different patient groups were created using simple linear regression.

Median-split of patient survival after TSPO-PET imaging was performed to distinguish between patients with long and short survival and an unpaired Student’s t-test served to test for significance between TSPO-PET signals of both groups. Median-split was used to define patients with high (> Median) and low (< Median) contralateral TSPO-PET signal. The survival period was defined as the interval between date of TSPO-PET and date of death. Univariate Cox regression and Log-rank test was performed to determine the impact of a battery of common variables and contralateral TSPO-PET signal on patient survival. Contralateral TSPO-PET signals were z-transformed due to lack of normal distribution in the sample of n=31 patients with glioblastoma with available survival data. Multivariate Cox-regression was applied to compare survival between both patient groups including contralateral TSPO-PET signal, TSPO-PET signal of the tumor, T1 contrast enhancement volumes, age and subsequent radiotherapy and chemotherapy as variables. Multivariate Cox regression was additionally controlled for initiation of glucocorticoid medication prior to TSPO-PET to exclude confounding effects of steroids on TSPO expression. Furthermore MGMT was included due to significant impact on survival in an extended cohort (22). As a validation, a multivariate Cox regression was performed including all tested variables.

#### Determination of the cellular source of contralateral TSPO-PET signal alterations

Unpaired Student’s t-test was used to compare contralateral TSPO-PET signal in mice with syngeneic SB28 glioblastoma and sham injection. Voxel-wise regression was performed via statistical parametric mapping as described previously (78) to perform data driven correlation of tumor TSPO-PET signals as a seed with TSPO-PET signal in the whole mouse brain. One-way ANOVA served to compare single cell tracer uptake between SB28 glioblastoma, sham-injected and healthy control mice. Unpaired Student’s t-test was used for comparison of cell type coverage in immunofluorescence. One-way ANOVA was applied for differentiation of gene expression in qPCR between groups.

#### Immune phenotype characterization

For calculation of the volcano plot, FDR-correction was used for multiple comparisons with a threshold of 0.05. Comparison of gene expression between contralateral hemispheres of SB28 glioblastoma and sham-injected mice was performed using unpaired Student’s t-tests.

### Study approval

All patients have given written informed consent prior to participation. The study was approved by the local ethics committee (approval number 17-457). Twenty healthy controls were obtained from the interdisciplinary Alzheimer’s disease study “Activity of Cerebral Networks, Amyloid and Microglia in Aging and AD (ActiGliA)” and an observational study on various neuroinflammatory conditions (ethics-applications: 601-16, 17-569, 17-755; radiation protection applications: Z 5 – 22463/2 – 2015 – 006, Z 5 - 22464/2017-047-K-G). All animal experiments were performed in compliance with the National Guidelines for Animal Protection, Germany and with the approval of the regional animal committee (Government of Upper Bavaria) and overseen by a veterinarian. All animals were housed in a temperature- and humidity-controlled environment with a 12-h light–dark cycle, with free access to food (Ssniff) and water.

## Data Availability

All data needed to evaluate the conclusions of Fig. 1-5 are present in the paper and/or the Supplementary Materials. Imaging data will be shared in DICOM format upon reasonable request to the corresponding author.

## Funding

This project was partly funded by the Deutsche Forschungsgemeinschaft (DFG, German Research Foundation) (FOR 2858 project numbers 421887978 and 422188432 and Research Training Group GRK 2274). NLA is supported by a research grant of the Else Kröner-Fresenius-Stiftung. MB was funded by the Deutsche Forschungsgemeinschaft (DFG) under Germany’s Excellence Strategy within the framework of the Munich Cluster for Systems Neurology (EXC 2145 SyNergy – ID 390857198). SVK was supported by the Verein zur Förderung von Wissenschaft und Forschung an der Medizinischen Fakultät der LMU München (WiFoMed) and the Friedrich-Baur-Stiftung. AH received funding by the Bavarian Cancer Research Center (BZKF) and is currently funded by the Deutsche Forschungsgemeinschaft (DFG, German Research Foundation) – 545058105.

## Author contributions

LMB: performed patient selection and human PET image analysis and interpretation; performed tumor inoculation and murine PET acquisition, image analysis and interpretation; performed scRadiotracing and analyzed scRadiotracing data; wrote the first draft of the manuscript with input of all co-authors. SQ, SVK, and JB: gathering clinical information about patients, interpretation of PET data in the context of clinical presentation of human glioblastoma. VZ: performed immunofluorescence, multiplex gene expression and qPCR analyses, interpretated histological data and contributed representative images. KWM, AH, LHK: participated in preclinical PET acquisition, image analysis and interpretation. LHK, ZIK and SU: participated in image analysis and interpretation of immunofluorescence data. VCR, JH: neuropathological examination and characterization of the tumor samples. STK, LH, MH and HEP: participated in tumor inoculation and scRadiotracing experiments; performed transcardial perfusion. MG, NF: contributed to data visualization. AZa, AZo, LK: performed PET image reconstruction and connectivity analyses. RP: contributed patient data of the healthy control group. BSR: contributed high resolution MRI data. MP, ST: interpretation of qPCR and multiplex gene analyses; pathway analyses. MJR, RP, SS, SZ, AE, JH, JCT, NT, LvB, MP, ST, NLA and MB: conception and design, contributed to interpreting data, enhancing intellectual content of manuscript. All authors contributed with intellectual content and revised the manuscript.

## Acknowledgements

We thank Rosel Oos and Giovanna Palumbo for excellent technical support during small animal PET imaging.

## Supplement

**Supplementary Table 1:** Univariate survival analysis. Univariate Cox regression was used to test for significance of distinct parameters and indices on patient survival. SNP = single nucleotide polymorphism. LAB = ow, MAB = medium, HAB = high affinity binding status. MGMT = O-6-Methylguanine-DNA methyltransferase. TERT = Telomerase reverse transcriptase. CE = contrast enhancement. *time indicated in months. **bold font highlights statistically significant associations.

|  | **Median overall survival (range)\*** | **Significance\*\*** |
| --- | --- | --- |
| **Age** |  | **p = 0.015** |
| Median < 72.6y, n=16 | 14.28 (1.50-57.17) |  |
| Median > 72.6y, n=15 | 6.53 (0.53-21.37) |  |
| **Sex** |  | **p = 0.953** |
| Male, n=21 | 10.83 (1.50-57.17) |  |
| Female, n=10 | 11.40 (0.53-43.73) |  |
| **SNP** |  | **p = 0.777** |
| LAB, n=5 | 10.53 (4.17-19.93) |  |
| MAB, n=16 | 11.58 (0.53-57.17) |  |
| HAB, n=10 | 7.17 (0.93-27.67) |  |
| **MGMT promoter methylation status** |  | **p = 0.581** |
| Methylated, n=13 | 11.57 (0.53-57.17) |  |
| Unmethylated, n=18 | 10.43 (0.93-27.67) |  |
| **TERT promoter mutation status** |  | **p = 0.352** |
| mutant, n=26 | 10.83 (0.53-43.73) |  |
| wildtype, n=4 | 19.43 (10.53-57.17) |  |
| **Glucocorticoid therapy** |  | **p = 0.053** |
| yes, n=11 | 5.17 (0.53-25.77) |  |
| no, n=20 | 11.58 (3.07-57.17) |  |
| **Tumor resection** |  | **p = 0.152** |
| yes, n=7 | 19.07 (1.50-57.17) |  |
| no, n=24 | 10.28 (0.53-43.73) |  |
| **Radiotherapy** |  | **p = 0.024** |
| yes, n=27 | 11.60 (0.93-57.17) |  |
| no, n=4 | 5.30 (0.53-11.57) |  |
| **Chemotherapy** |  | **p = 0.004** |
| yes, n=20 | 13.88 (4.03-57.17) |  |
| no, n=11 | 5.17 (0.53-25.77) |  |
| **Temozolomide** |  | **p = 0.306** |
| yes, n=14 | 13.40 (4.17-57.17) |  |
| no, n=17 | 9.10 (0.53-43.73) |  |
| **Tumor CE volume (cm3)** |  | **p < 0.001** |
| Median < 8.0, n=16 | 12.65 (4.17 - 57.17) |  |
| Median > 8.0, n=15 | 6.53 (0.53 - 27.67) |  |
| **Tumor T2 volume (cm3)** |  | **p < 0.001** |
| Median < 60.1, n=16 | 15.34 (0.53 - 57.17) |  |
| Median > 60.1, n=15 | 9.10 (0.93 - 21.37) |  |
| **Tumor TSPO-PET signal (SUVr)** |  | **p = 0.024** |
| Median < 2.5, n=16 | 13.58 (0.53 - 57.17) |  |
| Median > 2.5, n=15 | 9.10 (0.93 - 27.67) |  |
| **Contralateral TSPO-PET signal (z score)** |  | **p < 0.001** |
| Median < 0.5, n=16 | 13.40 (0.53 - 57.17) |  |
| Median > 0.5, n=15 | 6.53 (0.93 - 27.67) |  |

**Supplementary Table 2:** Multivariate survival analysis. Multivariate Cox regression was used to test for significance of relevant (significant in univariate Cox regression) parameters and indices on patient survival. Glucocorticoid medication was additionally included due to borderline significance. CE = contrast enhancement. MGMT = O-6-Methylguanine-DNA methyltransferase. MGMT was additionally included due to significance in an extended cohort. *bold font highlights statistically significant associations.

|  | **Hazard ratio (95% CI)** | **Significance\*** |
| --- | --- | --- |
| **Age** | 1.123 (1.055 - 1.196) | **p < 0.001** |
| **MGMT promoter methylation status** | 0.199 (0.058 - 0.684) | **p = 0.010** |
| **Glucocorticoid medication** | 5.162 (1.450 - 18.370) | **p = 0.011** |
| **Radiotherapy** | 0.274 (0.059 - 1.286) | p = 0.101 |
| **Chemotherapy** | 0.689 (0.251 - 1.896) | p = 0.471 |
| **Tumor CE volumes (cm3)** | 1.039 (1.005 - 1.074) | **p = 0.025** |
| **Tumor TSPO-PET signal (SUVr)** | 0.709 (0.286 - 1.760) | p = 0.459 |
| **Contralateral TSPO-PET signal (z score)** | 2.175 (1.263 - 3.744) | **p = 0.005** |

**Supplementary Table 3:** Extended multivariate survival analysis. Multivariate Cox regression was used to test for significance of all considered parameters and indices on patient survival. CE = contrast enhancement. MGMT = O-6-Methylguanine-DNA methyltransferase. *bold font highlights statistically significant associations.

|  | **Hazard ratio (95% CI)** | **Significance\*** |
| --- | --- | --- |
| **Age** | 1.204 (1.080 - 1.342) | **p < 0.001** |
| **Sex** | 1.111 (0.261 - 4.693) | p = 0.886 |
| **SNP** | 0.295 (0.089 - 0.984) | **p = 0.047** |
| **MGMT promoter methylation status** | 0.017 (0.001 - 0.196) | **p = 0.001** |
| **TERT promoter mutation status** | 0.421 (0.134 - 1.318) | p = 0.137 |
| **Glucocorticoid therapy** | 14.135 (1.595 - 125.302) | **p = 0.017** |
| **Tumor resection** | 0.253 (0.051 - 1.268) | p = 0.095 |
| **Radiotherapy** | 0.086 (0.007 - 1.088) | p = 0.058 |
| **Chemotherapy** | 1.215 (0.241 - 6.122) | p = 0.814 |
| **Tumor CE volumes (cm3)** | 1.061 (1.010 - 1.115) | **p = 0.019** |
| **Tumor T2 volumes (cm3)** | 1.004 (0.979 - 1.030) | p = 0.757 |
| **Tumor TSPO-PET signal (SUVr)** | 0.378 (0.100 - 1.428) | p = 0.151 |
| **Contralateral TSPO-PET signal (z score)** | 2.010 (1.105 - 3.656) | **p = 0.022** |

## Notes

### Author Declarations

The ethics committee of the Ludwig-Maximilians-University gave ethical approval for this work (approval number 17-457, approval numbers for data on healthy individuals 601-16, 17-569, 17-755)

### Summary of Updates

Typo in authors list corrected

